# Course and predictors of somatic symptom disorder in irritable bowel syndrome and ulcerative colitis: A longitudinal analysis from the SOMA.GUT-RCT

**DOI:** 10.64898/2025.12.01.25341343

**Authors:** Luisa Peters, Anna Matysiak, Sina Hübener, Ansgar W. Lohse, Bernd Löwe, Kerstin Maehder

**Affiliations:** Department of Psychosomatic Medicine and Psychotherapy, University Medical Center Hamburg-Eppendorf; Department of Medicine I, University Medical Center Hamburg-Eppendorf, Hamburg, Germany

**Keywords:** Somatic Symptom Disorder, Irritable Bowel Syndrome, Ulcerative Colitis, Diagnosis, Predictors, Course, Persistent Somatic Symptoms

## Abstract

**Background:** Longitudinal data on the course of somatic symptom disorder (SSD) in ulcerative colitis (UC) and irritable bowel syndrome (IBS) are lacking. Understanding SSD trajectories and predictors in IBS and UC may clarify clinical relevance and guide psychological treatment decisions. This study examined the 12-month course and biopsychosocial predictors of interview-based SSD in patients with UC or IBS.

**Methods:** Longitudinal data from a randomised controlled trial were analysed. SSD was assessed using DSM-5-based structured interviews at baseline and 12 months. SSD Criteria A (Somatic symptom severity) and B (symptom-related distress) were measured with the Patient Health Questionnaire-15 (PHQ-15) and the Somatic Symptom Disorder – B Criteria Scale-12 (SSD-12), respectively. Further variables included gastrointestinal symptom severity, inflammatory markers, depression severity, illness perceptions, and neuroticism. Logistic and linear regression models identified baseline predictors of SSD diagnosis and Criteria A and B at 12 months.

**Results:** The sample included 213 patients (73.7% female; *M*_age_=40.5, *SD*=13.98) with UC (*n*=110) or IBS (*n*=103). SSD was present in 42.3% (95%CI: 35.2–49.3) at baseline and in 15.5% (95%CI: 11.3– 20.7) at follow-up. Baseline SSD and depression severity predicted follow-up SSD. Criterion A was predicted by somatic symptom severity, female gender, and neuroticism; the B criterion by symptom-related distress, somatic symptom severity, neuroticism, and negative illness perceptions. Inflammatory markers and gastrointestinal symptom severity showed no predictive value.

**Conclusion:** Structured interview-based SSD was frequent in patients with UC or IBS at baseline and declined over time. Psychosocial rather than disease-related variables predicted SSD, highlighting modifiable targets for early detection and tailored interventions. Results should be interpreted in light of the study’s interventional context.

## Introduction

Somatic symptom disorder (SSD), as defined in the Diagnostic and Statistical Manual of Mental Disorders, Fifth Edition (DSM-5), is characterised by distressing somatic symptoms persisting for at least six months and excessive symptom-related thoughts, feelings, or behaviour (1). In contrast to earlier concepts focused on medically unexplained symptoms, SSD emphasises psychological distress associated with somatic complaints regardless of biomedical aetiology (2). This has increased the diagnosis’s clinical applicability across medical conditions, including gastrointestinal diseases. Ulcerative colitis (UC) and irritable bowel syndrome (IBS) are two chronic gastrointestinal conditions with high symptom burden and reduced quality of life (3, 4). While UC is an inflammatory bowel disease involving immune-mediated inflammation (5), IBS is considered a disorder of gut-brain interaction (6). Despite different aetiologies, both conditions involve persistent gastrointestinal symptoms such as abdominal pain and altered bowel habits (7), and are associated with psychological distress, anxiety and depression (8, 9). Symptoms in UC often persist without active inflammation (10), while in IBS they are one diagnostic feature (11). Psychological processes like illness-related anxiety, hypervigilance, and catastrophising influence symptom experience and illness behaviour via bidirectional gut-brain interactions (12, 13). These factors also predicted disease-related outcomes such as disease onset or activity in both conditions (14-16). Despite the frequent occurrence of persistent symptoms and the known impact of psychological processes, SSD has rarely been studied in IBS or UC (17, 18).

Most studies on SSD rely on self-report measures, which may overestimate prevalence (17), while structured diagnostic interviews remain underused in medical conditions like UC and IBS (17). In a cross-sectional investigation of the same sample used in the present study, 55% of patients with IBS and 29.6% with UC met diagnostic criteria for SSD at baseline (19). As these data are drawn from a randomised controlled trial (RCT) targeting individuals with moderate symptoms and interest in psychological support, these figures likely overestimate SSD frequency. Still, the observed frequencies underscore SSD’s clinical relevance in both conditions. This questions the International Classification of Diseases, 11th Revision (ICD-11) exclusion of bodily distress disorder (BDD), the diagnostic equivalent of SSD, from IBS, especially given limited empirical support for this criterion (20).

To date, no study has prospectively examined the course and predictors of SSD in patients with IBS or UC using structured diagnostic interviews. (17, 18). Understanding this course is essential to evaluate diagnostic relevance in these two conditions, identify those who need psychological care, and guide timely interventions. Longitudinal data from a psychosomatic outpatient sample suggest that SSD often follows a persistent course: After four years, over 30% of patients with SSD at baseline still met diagnostic criteria, while only 21% had remitted (21). High symptom-related distress and elevated anxiety severity at baseline predicted SSD persistence, whereas somatic symptom severity alone did not (21). A recent meta-analysis (18) further identified anxiety, depression, and a self-concept of bodily weakness as relevant predictors of persistent SSD, while also highlighting the overall scarcity and inconsistency of longitudinal data on SSD.

An SSD diagnosis in UC or IBS may offer clinical value by identifying individuals with high symptom-related distress and guiding decisions about when to integrate psychological treatment alongside biomedical care. In this context, it is essential to understand how SSD develops in these patient groups and which factors influence its course. To address this, the present study investigated the course and predictors of SSD in patients with IBS or UC using structured diagnostic interviews. We assessed the frequency of SSD at baseline and 12-month follow-up, as well as the frequency of specific SSD trajectories (persistence, remission, new cases, no SSD). Additionally, we explored sociodemographic, psychosocial, and disease-related predictors of SSD diagnosis at follow-up. To complement this categorical outcome, we also examined SSD criteria A (somatic symptom severity) and B (excessive symptom-related distress) as dimensional outcomes and explored their respective predictors.

## Methods

### Study design

This longitudinal study was conducted as part of the three-arm RCT SOMA.GUT, which evaluated a mechanism-based psychological intervention targeting dysfunctional symptom expectations and illness-related anxiety in patients with IBS or UC. Patients were randomised (1:1:1) to (1) the mechanism-based intervention (GUT.EXPECT⍰+⍰standard care), (2) a non-specific supportive intervention (GUT.SUPPORT⍰+⍰standard care), or (3) standard care (SC) only. The trial was registered (ISRCTN30800023) and has been completed; full details are available in the published study protocol (22). Primary and relevant secondary outcomes of the trial have been reported previously (23). No between-group differences in gastrointestinal symptom severity were observed at the primary endpoint at 3 months. Exploratory analyses at 12 months suggest greater improvements in gastrointestinal symptom severity in GUT.EXPECT + SC compared to SC only. No differential treatment effects were observed for UC and IBS (23). Based on these findings, the present study examined SSD frequency, trajectories and predictors across the full sample, considering trial arm in analyses. SOMA.GUT is part of the DFG-funded (Deutsche Forschungsgemeinschaft) research unit RU 5211 SOMACROSS, investigating biopsychosocial mechanisms of symptom persistence across medical conditions (24). Ethical approval for the SOMA.GUT trial was granted by the Ethics Committee of the Hamburg Medical Association (reference number: 2020-10198-BO-ff).

### Patient recruitment and procedures

Participants were recruited nationwide between April 2022 and February 2024 as part of the SOMA.GUT trial. Recruitment was coordinated by the Department of Psychosomatic Medicine and Psychotherapy at the University Medical Center Hamburg-Eppendorf. Details are in the primary publication of the trial (23). Eligible individuals were ≥18 years old, fluent in German, and diagnosed with UC or IBS, confirmed by medical records reviewed by a study physician; IBS also required fulfilment of Rome⍰IV criteria (11). Inclusion also required moderate or greater gastrointestinal symptom severity (IBS-Severity Scoring System score ≥175 (25)), and ongoing treatment in accordance with national guidelines (26, 27). Exclusion criteria included medical emergencies, acute suicidality, florid psychosis, or recent psychotherapy (past 3 months). After informed consent, participants were randomised to one of three study arms. At baseline, they completed questionnaires and provided blood and stool samples. SSD was assessed via structured diagnostic telephone interview and reassessed after⍰12⍰months. Participants in the intervention arms received four⍰45⍰minute videoconference sessions over three months; all groups continued standard medical care. Analyses include participants with complete SSD data at both time points.

### Instruments

#### Assessment of SSD

SSD was assessed at baseline and 12-month follow-up using semi-structured diagnostic interviews based on the SSD section of the Structured Clinical Interview for DSM-5 (research version; with permission from the American Psychiatric Association) (28, 29). SSD severity was classified according to DSM-5 criteria as mild (1 criterion B fulfilled), moderate (≥2 criteria B fulfilled), or severe (≥2 criteria B fulfilled plus multiple somatic complaints or one particularly severe symptom) (1). Telephone interviews were conducted by six trained research assistants with medical or psychological backgrounds, lasted 15–45 minutes, and were reviewed by a clinical supervisor. To complement the interviews, SSD Criteria A and B were also assessed via self-report. Criterion A was measured via somatic symptom severity assessed with the Patient Health Questionnaire-15 (PHQ-15; range 0–30) (30), which differentiates minimal (0–4), low (5–9), medium (10–14), and high (15–30) symptom severity, and shows high internal consistency and good construct validity (31). Criterion B was assessed via the Somatic Symptom Disorder – B Criteria Scale-12 (SSD-12; range 0–48) (32) which measures cognitive, affective, and behavioral aspects of symptom-related distress and has demonstrated high internal consistency and good construct validity (33).

#### Sociodemographic and psychosocial variables

Sociodemographic variables included gender, age, educational level, employment status, and marital status. The following psychosocial variables were assessed:

Depression severity was measured using the Patient Health Questionnaire-9 (PHQ-9; range 0–27) (34, 35) and anxiety severity was assessed via the Generalized Anxiety Disorder Scale-7 (GAD-7; range 0– 21) (36, 37), both are well-validated screening tools (34, 36). Illness perceptions were assessed with the Brief Illness Perception Questionnaire (B-IPQ; range 0–80) (38); demonstrating good reliability and validity (39, 40). Perceived stress was measured with the Perceived Stress Scale-10 (PSS-10; range 0–40) (41), showing high internal consistency and factorial validity (42). Alexithymia was assessed with the Toronto Alexithymia Scale-20 (TAS-20; range 20–100) (43), which has strong internal consistency and test-retest reliability (44). Adverse childhood experiences were evaluated with the Adverse Childhood Experiences Questionnaire (ACE-D; range 0–10) (45), a reliable and economical retrospective screening of adverse childhood experiences. Neuroticism was measured with the neuroticism subscale of the Big Five Inventory-10 (BFI-10; range 2–10, higher scores reflect lower neuroticism) (46), which shows acceptable internal consistency and good convergent validity (47).

#### Disease-related outcomes

Disease-related variables included time since diagnosis and symptom onset, both assessed via self-report. For UC, disease activity was measured with a single-item flare-up question (yes/no) and the Simple Clinical Colitis Activity Index (SCCAI; range 0–19) (48). The SCCAI covers bowel frequency (day/night), urgency, stool blood, general well-being, and extraintestinal manifestations, and correlates well with more complex indices (48). Inflammatory activity was assessed with C-reactive protein (CRP; ≥5 mg/L = elevated) and faecal calprotectin (≥50 μg/g = elevated). Gastrointestinal symptom severity was measured using the Irritable Bowel Syndrome – Severity Scoring System (IBS-SSS; range 0–500) (25), which assesses abdominal pain, bloating, dissatisfaction with bowel habits, and bowel-related impairment over the past 10 days. Severity is categorised as mild (75–174), moderate (175–299), or severe (≥300). Originally designed for IBS, the IBS-SSS is also applied in inflammatory bowel disease and is sensitive to changes in gastrointestinal symptom severity (49, 50).

### Statistical analysis

All analyses were conducted using IBM SPSS Statistics for Windows, Version 29.0 (IBM Corp., Armonk, NY, USA). Missing data were minimal: seven participants were excluded from the respective analyses involving the B-IPQ and one from the TAS-20 due to incomplete responses. Descriptive statistics summarised sample characteristics and SSD frequencies overall and by diagnosis at baseline and follow-up. Chi-square tests compared SSD frequency between bowel disease type and trial arms at both time point, and SSD severity distribution over time. Participants were categorised into four SSD trajectory groups: (1) persistent SSD (diagnosis at both time points), (2) remitted SSD (baseline only), (3) new SSD (follow-up only), and (4) no SSD (neither time point). Differences in the distribution of SSD trajectory groups were examined across bowel disease type, trial arms, sociodemographic variables as well as disease-related categorical variables using chi-square tests. Continuous disease-related variables were compared between SSD trajectory groups using one-way analyses of variance. Predictors of SSD diagnosis at follow-up were analysed using hierarchical logistic regression following a bio-psycho-social approach. Step 1 included trial arm allocation (intervention group vs. standard care), baseline SSD diagnosis, and sociodemographic factors including age, gender, and education. Step 2 added somatic symptom severity (PHQ-15) as predictor for SSD criterion A and further disease-related factors, including bowel disease type (IBS/UC), baseline gastrointestinal symptom severity (IBS-SSS), and inflammation markers (CRP, faecal calprotectin). Step 3 introduced symptom-related distress (SSD-12) as predictor for SSD B criteria, and further psychosocial factors, including depression severity (PHQ-9), anxiety severity (GAD-7), perceived stress (PSS-10), neuroticism (BFI-10), childhood adversity (ACE-D), alexithymia (TAS-20), and illness perceptions (B-IPQ). No multicollinearity was detected (Pearson’s *r* < .70). Results are reported as odds ratios with 95% confidence intervals, *p*-values, and Nagelkerke’s R^2^. Two hierarchical linear regressions explored predictors of SSD Criterion A (somatic symptom severity, PHQ-15) and B (symptom-related distress, SSD-12). Step 1 included trial arm allocation and the baseline score of the respective outcome (PHQ-15 or SSD-12). Step 2 added disease-related factors while step 3 introduced psychosocial factors. Multicollinearity was checked via variance inflation factors (VIF), all within acceptable limits. Results are presented as unstandardised coefficients (*b*), standard errors *(SE*), standardised betas (β), and *p*-values. Adjusted *R*^2^ values were used to indicate model fit. No correction for multiple testing was applied given the exploratory aim.

## Results

### Study sample

Of the 247 patients enrolled in the SOMA.GUT-RCT, 11 were excluded at baseline due to major protocol violations (*n* = 5), incomplete baseline data (*n* = 3), withdrawal of consent (*n* = 2), or missing SSD interview data (*n* = 1), resulting in a baseline sample of 236 participants. At the 12-month follow-up, 23 additional participants were excluded (study withdrawal *n* = 6, no further data provided *n* = 5, unreachable for diagnostic interview *n* = 12), resulting in a final sample of *N* = 213 patients with UC (*n* = 110) or IBS (*n* = 103). The mean age was 40.5 years (*SD* = 13.98), 45.5% held a university degree, and 73.7% identified as female (see **Table 1** for more details). Compared to participants included in the final analysis, those lost to follow-up reported higher baseline gastrointestinal symptom severity (*p* = 0.022); all other characteristics were comparable (see **Supplement 1**).

**Table 1.**
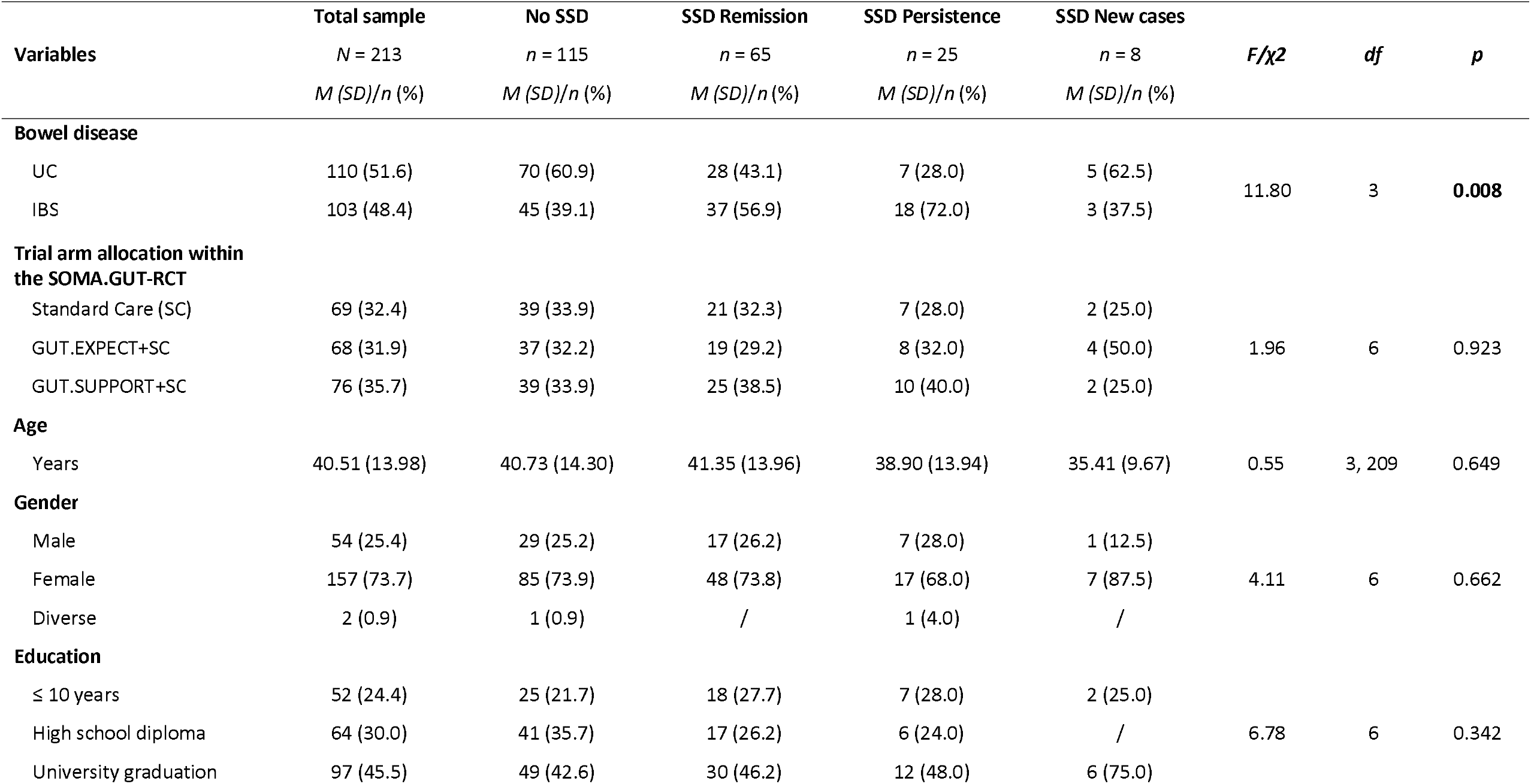
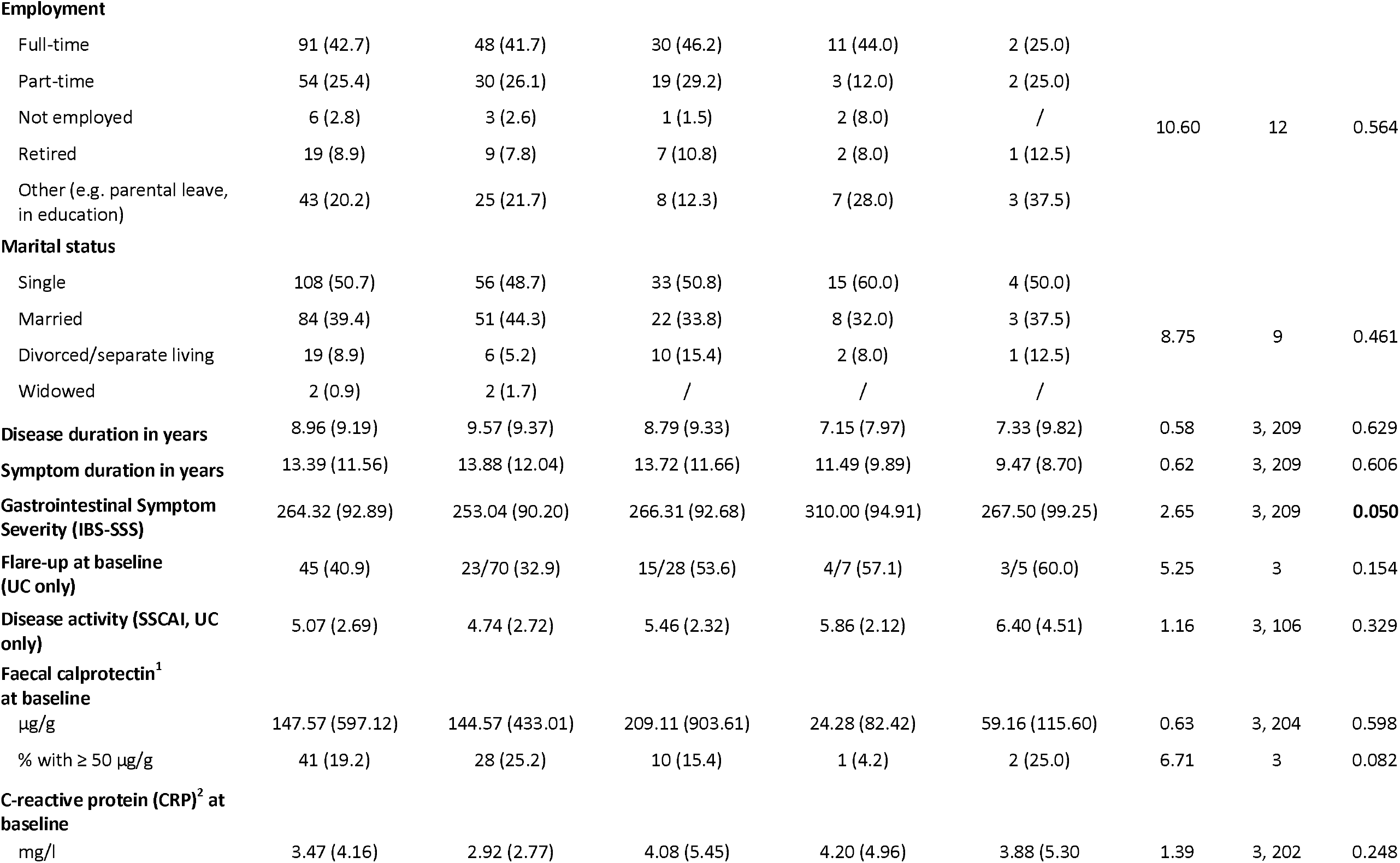
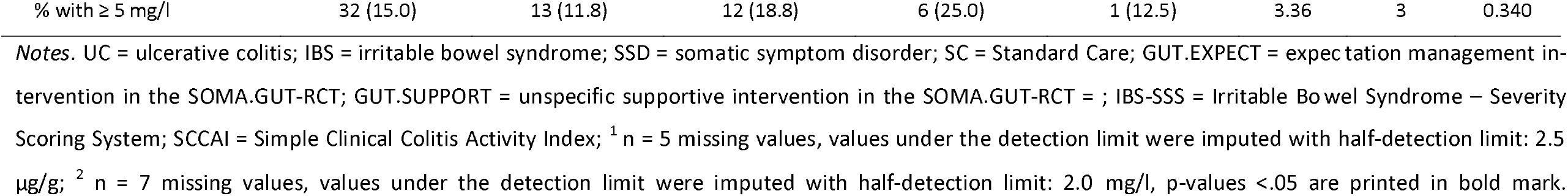
Baseline sociodemographic and disease-related characteristics of the total sample and the SSD trajectory groups No SSD, SSD Remission, SSD Persistence and SSD New cases

| **Variables** | **Total sample**<br>*N* = 213<br>*M (SD)/n (%)* | **No SSD**<br>*n* = 115<br>*M (SD)/n (%)* | **SSD Remission**<br>*n* = 65<br>*M (SD)/n (%)* | **SSD Persistence**<br>*n* = 25<br>*M (SD)/n (%)* | **SSD New cases**<br>*n* = 8<br>*M (SD)/n (%)* | *F/χ<sup>2</sup>* | *df* | *p* |
| --- | --- | --- | --- | --- | --- | --- | --- | --- |
| **Bowel disease** |  |  |  |  |  |  |  |  |
| UC | 110 (51.6) | 70 (60.9) | 28 (43.1) | 7 (28.0) | 5 (62.5) | 11.80 | 3 | **0.008** |
| IBS | 103 (48.4) | 45 (39.1) | 37 (56.9) | 18 (72.0) | 3 (37.5) |  |  |  |
| **Trial arm allocation within the SOMA.GUT-RCT** |  |  |  |  |  |  |  |  |
| Standard Care (SC) | 69 (32.4) | 39 (33.9) | 21 (32.3) | 7 (28.0) | 2 (25.0) | 1.96 | 6 | 0.923 |
| GUT.EXPECT+SC | 68 (31.9) | 37 (32.2) | 19 (29.2) | 8 (32.0) | 4 (50.0) |  |  |  |
| GUT.SUPPORT+SC | 76 (35.7) | 39 (33.9) | 25 (38.5) | 10 (40.0) | 2 (25.0) |  |  |  |
| **Age** |  |  |  |  |  |  |  |  |
| Years | 40.51 (13.98) | 40.73 (14.30) | 41.35 (13.96) | 38.90 (13.94) | 35.41 (9.67) | 0.55 | 3, 209 | 0.649 |
| **Gender** |  |  |  |  |  |  |  |  |
| Male | 54 (25.4) | 29 (25.2) | 17 (26.2) | 7 (28.0) | 1 (12.5) | 4.11 | 6 | 0.662 |
| Female | 157 (73.7) | 85 (73.9) | 48 (73.8) | 17 (68.0) | 7 (87.5) |  |  |  |
| Diverse | 2 (0.9) | 1 (0.9) | / | 1 (4.0) | / |  |  |  |
| **Education** |  |  |  |  |  |  |  |  |
| ≤ 10 years | 52 (24.4) | 25 (21.7) | 18 (27.7) | 7 (28.0) | 2 (25.0) | 6.78 | 6 | 0.342 |
| High school diploma | 64 (30.0) | 41 (35.7) | 17 (26.2) | 6 (24.0) | / |  |  |  |
| University graduation | 97 (45.5) | 49 (42.6) | 30 (46.2) | 12 (48.0) | 6 (75.0) |  |  |  |
| **Employment** |  |  |  |  |  |  |  |  |
| Full-time | 91 (42.7) | 48 (41.7) | 30 (46.2) | 11 (44.0) | 2 (25.0) |  |  |  |
| Part-time | 54 (25.4) | 30 (26.1) | 19 (29.2) | 3 (12.0) | 2 (25.0) |  |  |  |
| Not employed | 6 (2.8) | 3 (2.6) | 1 (1.5) | 2 (8.0) | / | 10.60 | 12 | 0.564 |
| Retired | 19 (8.9) | 9 (7.8) | 7 (10.8) | 2 (8.0) | 1 (12.5) |  |  |  |
| Other (e.g. parental leave, in education) | 43 (20.2) | 25 (21.7) | 8 (12.3) | 7 (28.0) | 3 (37.5) |  |  |  |
| **Marital status** |  |  |  |  |  |  |  |  |
| Single | 108 (50.7) | 56 (48.7) | 33 (50.8) | 15 (60.0) | 4 (50.0) |  |  |  |
| Married | 84 (39.4) | 51 (44.3) | 22 (33.8) | 8 (32.0) | 3 (37.5) | 8.75 | 9 | 0.461 |
| Divorced/separate living | 19 (8.9) | 6 (5.2) | 10 (15.4) | 2 (8.0) | 1 (12.5) |  |  |  |
| Widowed | 2 (0.9) | 2 (1.7) | / | / | / |  |  |  |
| **Disease duration in years** | 8.96 (9.19) | 9.57 (9.37) | 8.79 (9.33) | 7.15 (7.97) | 7.33 (9.82) | 0.58 | 3, 209 | 0.629 |
| **Symptom duration in years** | 13.39 (11.56) | 13.88 (12.04) | 13.72 (11.66) | 11.49 (9.89) | 9.47 (8.70) | 0.62 | 3, 209 | 0.606 |
| **Gastrointestinal Symptom Severity (IBS-SSS)** | 264.32 (92.89) | 253.04 (90.20) | 266.31 (92.68) | 310.00 (94.91) | 267.50 (99.25) | 2.65 | 3, 209 | **0.050** |
| **Flare-up at baseline (UC only)** | 45 (40.9) | 23/70 (32.9) | 15/28 (53.6) | 4/7 (57.1) | 3/5 (60.0) | 5.25 | 3 | 0.154 |
| **Disease activity (SSCAI, UC only)** | 5.07 (2.69) | 4.74 (2.72) | 5.46 (2.32) | 5.86 (2.12) | 6.40 (4.51) | 1.16 | 3, 106 | 0.329 |
| **Faecal calprotectin<sup>1</sup> at baseline** |  |  |  |  |  |  |  |  |
| µg/g | 147.57 (597.12) | 144.57 (433.01) | 209.11 (903.61) | 24.28 (82.42) | 59.16 (115.60) | 0.63 | 3, 204 | 0.598 |
| % with ≥ 50 µg/g | 41 (19.2) | 28 (25.2) | 10 (15.4) | 1 (4.2) | 2 (25.0) | 6.71 | 3 | 0.082 |
| **C-reactive protein (CRP)<sup>2</sup> at baseline** |  |  |  |  |  |  |  |  |
| mg/l | 3.47 (4.16) | 2.92 (2.77) | 4.08 (5.45) | 4.20 (4.96) | 3.88 (5.30) | 1.39 | 3, 202 | 0.248 |
| % with $\geq 5$ mg/l | 32 (15.0) | 13 (11.8) | 12 (18.8) | 6 (25.0) | 1 (12.5) | 3.36 | 3 | 0.340 |

*Notes.* UC = ulcerative colitis; IBS = irritable bowel syndrome; SSD = somatic symptom disorder; SC = Standard Care; GUT.EXPECT = expectation management intervention in the SOMA.GUT-RCT; GUT.SUPPORT = unspecific supportive intervention in the SOMA.GUT-RCT = ; IBS-SSS = Irritable Bowel Syndrome – Severity Scoring System; SCCAI = Simple Clinical Colitis Activity Index; <sup>1</sup> n = 5 missing values, values under the detection limit were imputed with half-detection limit: 2.5 $\mu\text{g/g}$ ; <sup>2</sup> n = 7 missing values, values under the detection limit were imputed with half-detection limit: 2.0 mg/l, p-values <.05 are printed in bold mark.

### Frequencies of SSD at baseline and 12-month follow-up

**Figure 1** displays the frequency distribution of SSD at baseline and 12-month follow-up. At baseline, 42.3% of patients (90/213; 95% CI: 35.2–49.3) met diagnostic criteria of SSD, decreasing to 15.5% (33/213; 95% CI: 11.3–20.7) after 12 months (*χ*^2^(1) = 37.14, *p* < 0.001). In the IBS group, frequency dropped from 53.4% (55/103; 95% CI: 43.9–63.2) to 20.4% (21/103; 95% CI: 13.4–28.2) (*χ*^2^(1) = 24.10, *p* < 0.001). In UC, rates declined from 31.8% (35/110; 95% CI: 23.0–40.0) to 10.9% (12/110; 95% CI: 5.2–17.6) (*χ*^2^(1) = 14.31, *p* < 0.001). SSD was more frequent in IBS than UC at baseline (*χ*^2^(1) = 10.15, *p* = 0.001), with only a trend at follow-up (*χ*^2^(1) = 3.65, *p* = 0.056). SSD severity also shifted over time. Mild SSD increased from 54.4% to 78.8%, while moderate cases decreased from 38.9% to 18.2% and severe cases from 6.7% to 3.0% (*χ*^2^(2) = 6.01, *p* = 0.049). Across trial arms, SSD was present at baseline in 40.6% of patients in standard care (28/69), 39.7% in GUT.EXPECT + SC (27/68), and 46.1% in GUT.SUPPORT + SC (35/76) (*χ*^2^(2) = 0.71, *p* = 0.701). At follow-up, rates declined in all trial arms (SC: 13.0%; GUT.EXPECT + SC: 17.6%; GUT.SUPPORT + SC: 15.8%), with no between-group differences (*χ*^2^(2) = 0.56, *p* = 0.755).

**Figure 1.**
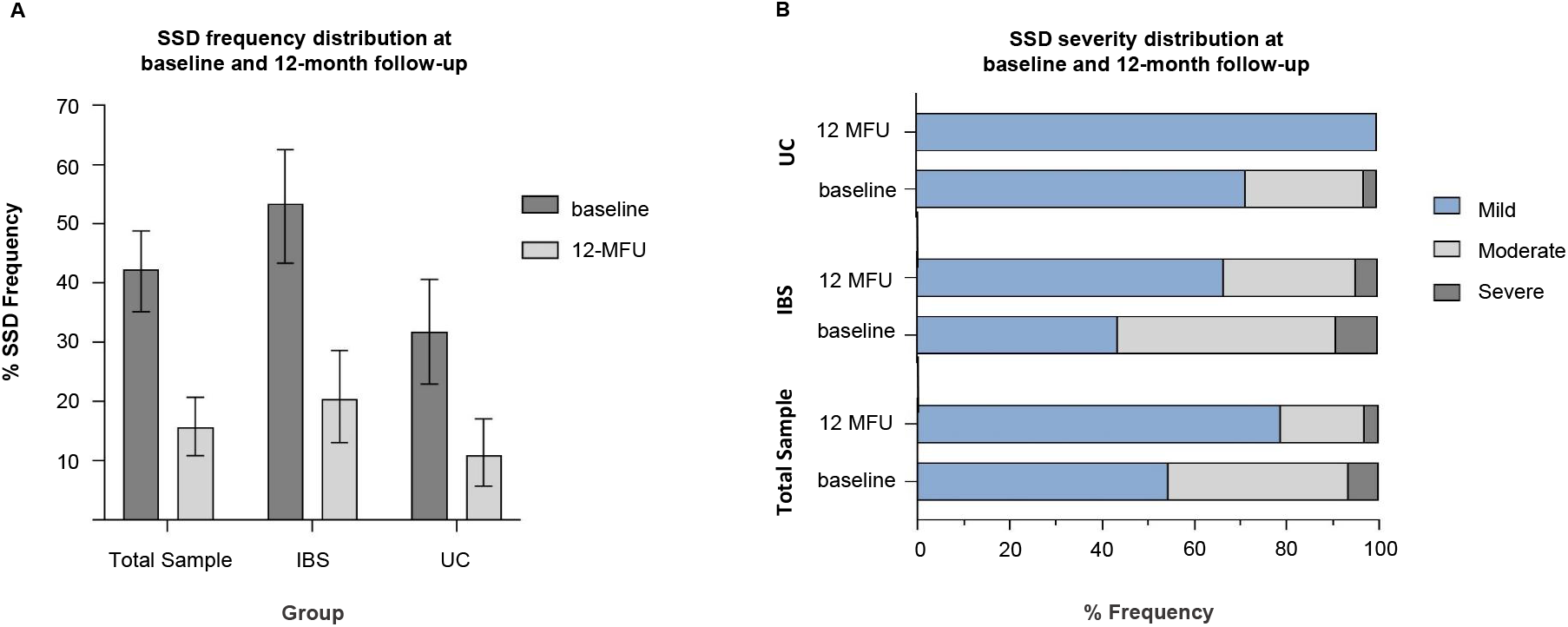
SSD frequency distribution at baseline and at 12-month follow-up (A) and SSD severity distribution at baseline and 12-month follow-up (B). **(A)** shows the proportion of patients with comorbid SSD in the total sample (*N* = 213), in patients with UC (*n* = 110) and patients with IBS (*n* = 103) at baseline and 12-month follow-up. Data are shown as percentage and 95% confidence intervals. **(B)** shows the severity distribution of SSD at baseline and 12-month follow-up within the total sample and in the UC and the IBS group respectively. Severity is divided into mild, moderate and severe. Data are shown as percentage and refer to 100% of patients with SSD in each group. SSD = somatic symptom disorder; UC = ulcerative colitis; IBS = irritable bowel syndrome; 12-MFU = 12-month follow-up assessment.

### Frequencies of SSD trajectories

**Figure 2** shows the distribution of SSD trajectories. Overall, 54.0% of patients had no SSD at either time point, 30.5% showed remission, 11.7% had persistent SSD, and 3.8% were newly diagnosed at follow-up. In the IBS group, 43.7% had no SSD, 35.9% remitted, 17.5% had persistent SSD, and 2.9% were newly diagnosed. In comparison, among patients with UC, 63.6% showed no SSD, 25.5% remitted, 6.4% had persistent SSD, and 4.5% were newly diagnosed at 12-month follow-up. The distribution of SSD trajectory groups differed between IBS and UC (χ^2^(3) = 11.80, *p* = 0.008), but not by trial arm or sociodemographic characteristics. Patients with persistent SSD reported significantly higher baseline gastrointestinal symptom severity than those without SSD (*p* = 0.028; see **Table 1**).

**Figure 2.**
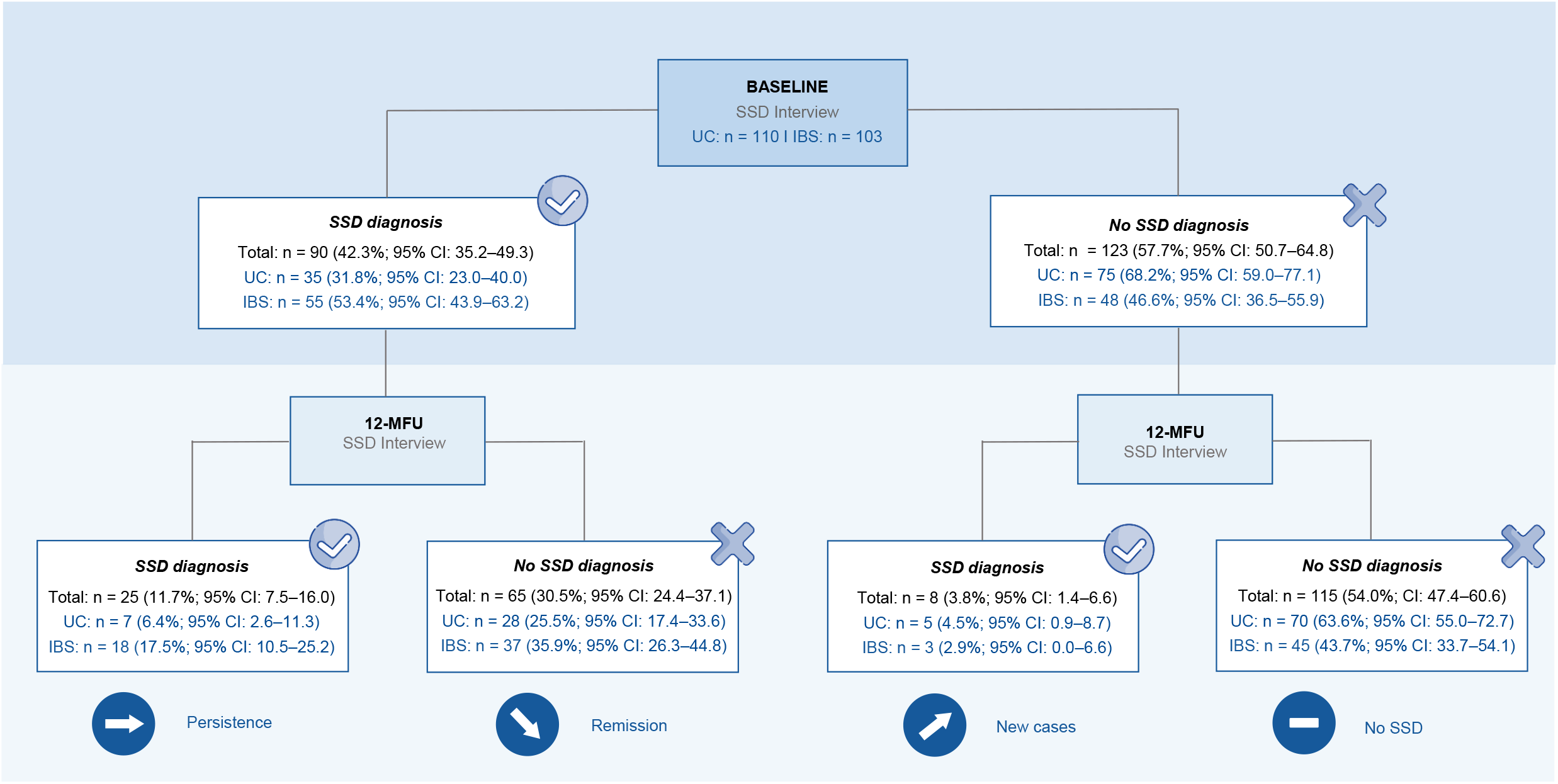
Number of patients fulfilling SSD diagnosis at baseline and 12-month follow-up, resulting in SSD trajectory groups *persistence, remission, new cases* and *no SSD* at 12-month follow-up in the total sample (*N* = 213), and in patients with UC (*n* = 110) and IBS (*n* = 103) separately. Results are presented as total number, percentage and 95% confidence interval. SSD = somatic symptom disorder, UC = ulcerative colitis; IBS = irritable bowel syndrome; CI = confidence interval; 12-MFU = 12-month follow-up assessment.

### Predictors of SSD diagnosis at 12-month follow-up

**Table 2** shows the results from the hierarchical logistic regression analysis predicting SSD diagnosis at 12-months. Baseline SSD was a significant predictor of follow-up SSD (*OR* = 3.51, *p* = 0.023). Higher baseline depression severity also predicted SSD at 12 months (*OR* = 1.21, *p* = 0.017). No other sociodemographic, disease-related, including gastrointestinal symptom severity or inflammatory activity, or psychosocial variables contributed to a relevant degree. The model explained a moderate proportion of variance (Nagelkerke’s *R*^2^ = 0.324) (see also **Supplement 2**).

**Table 2.**
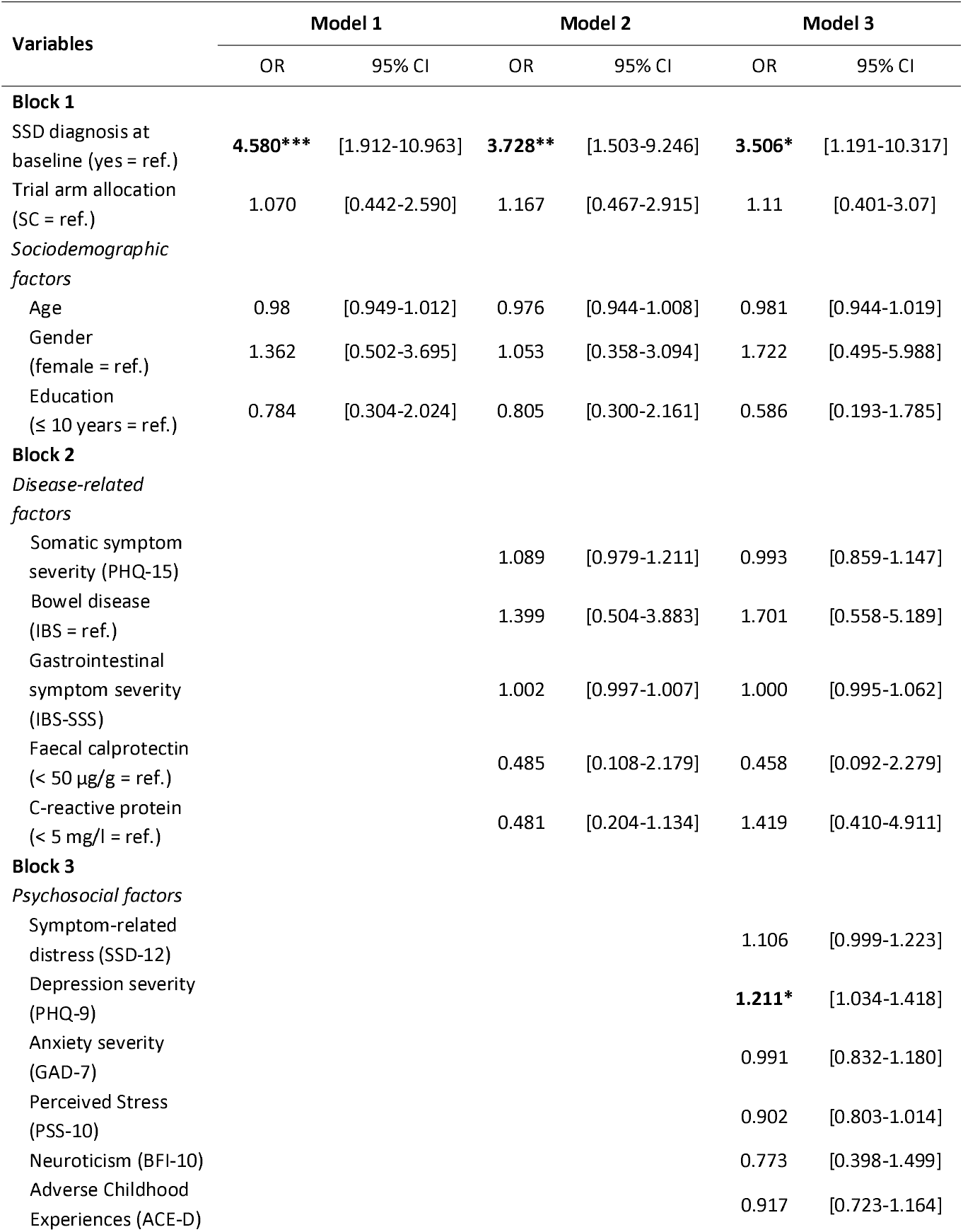
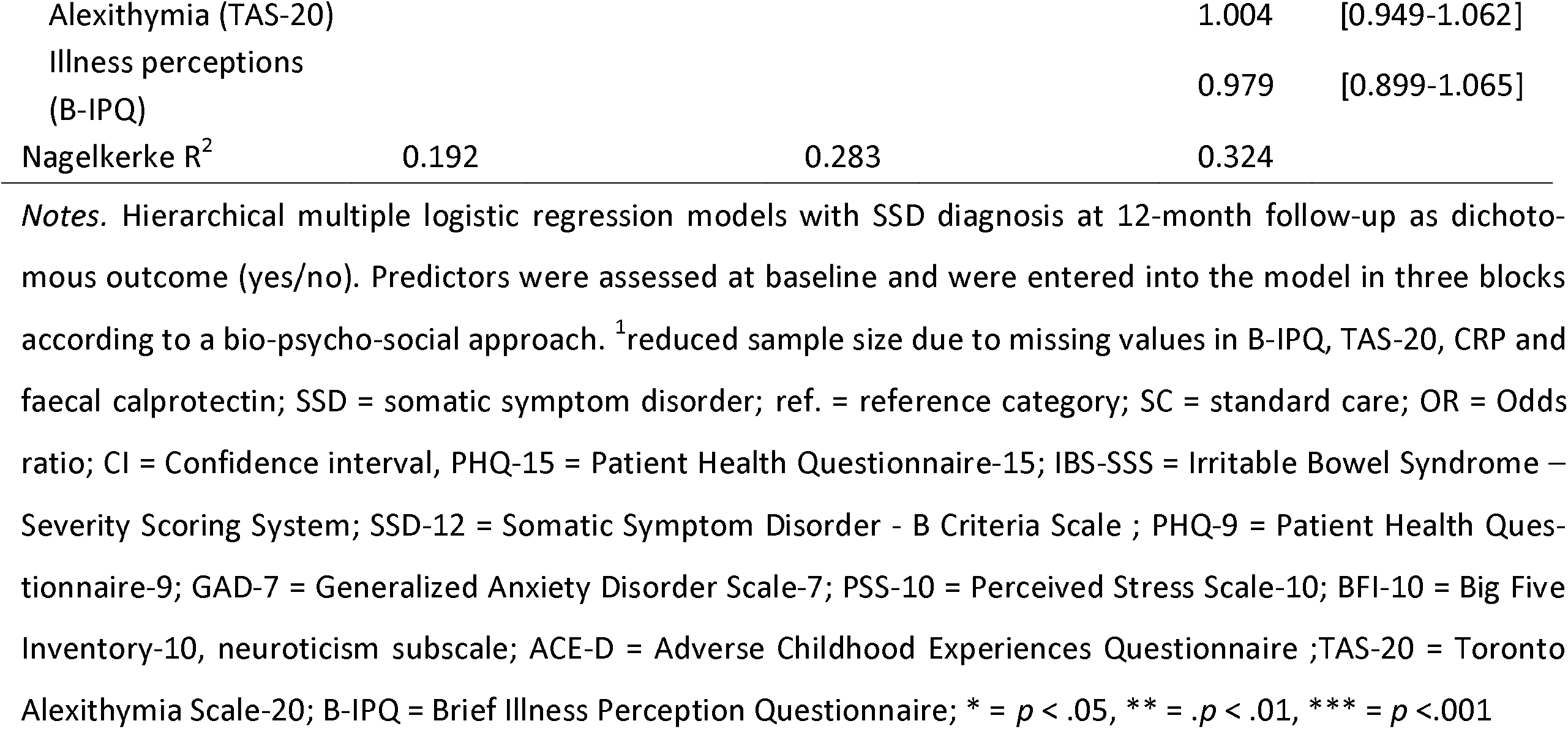
Hierarchical multiple logistic regression models to test baseline predictors of SSD diagnosis at 12-month follow-up in the total sample (N = 196^1^)

### Predictors of SSD A and B criteria at 12-month follow-up

Findings from the hierarchical linear regression models predicting self-reported SSD criteria A and B at 12-months are summarised in **Tables 3** and **Table 4**, respectively. For criterion A (somatic symptom severity, PHQ-15), the final model explained 41.6% (adjusted R^2^) of the variance at follow-up. Relevant predictors were higher baseline somatic symptom severity (*p* < 0.001), female gender (*p* = 0.025), and higher neuroticism (*p* = 0.004). No other variables contributed. For criterion B (symptom-related distress, SSD-12), the final model accounted for 43.7% (adjusted *R*^*2*^) of the variance. Baseline symptom-related distress (*p* < 0.001), somatic symptom severity (*p* = 0.023), higher neuroticism (*p* = 0.014), and more negative illness perceptions (*p* = 0.041) predicted greater symptom-related distress at follow-up (see also **Supplement 2**).

**Table 3.**
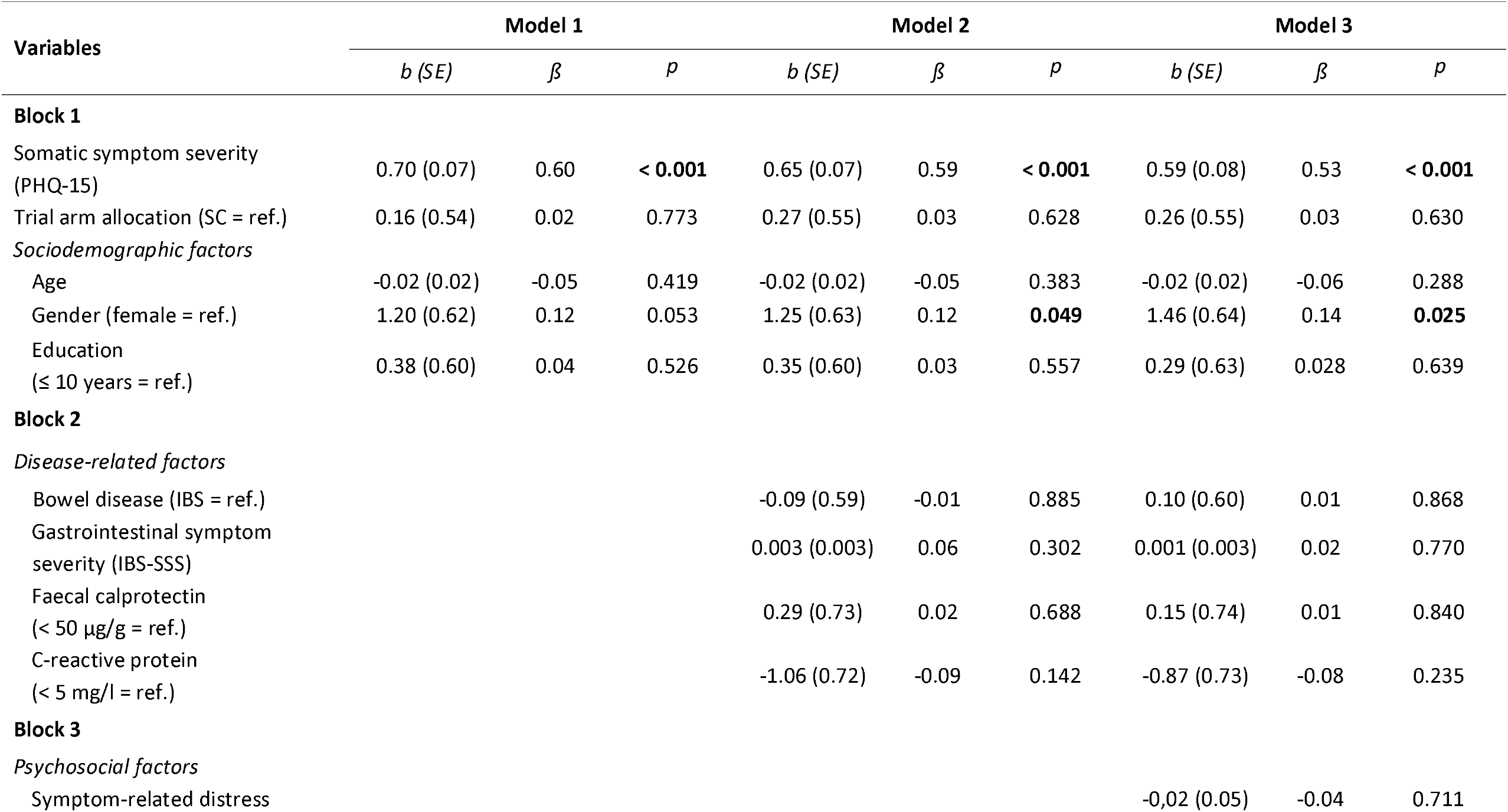
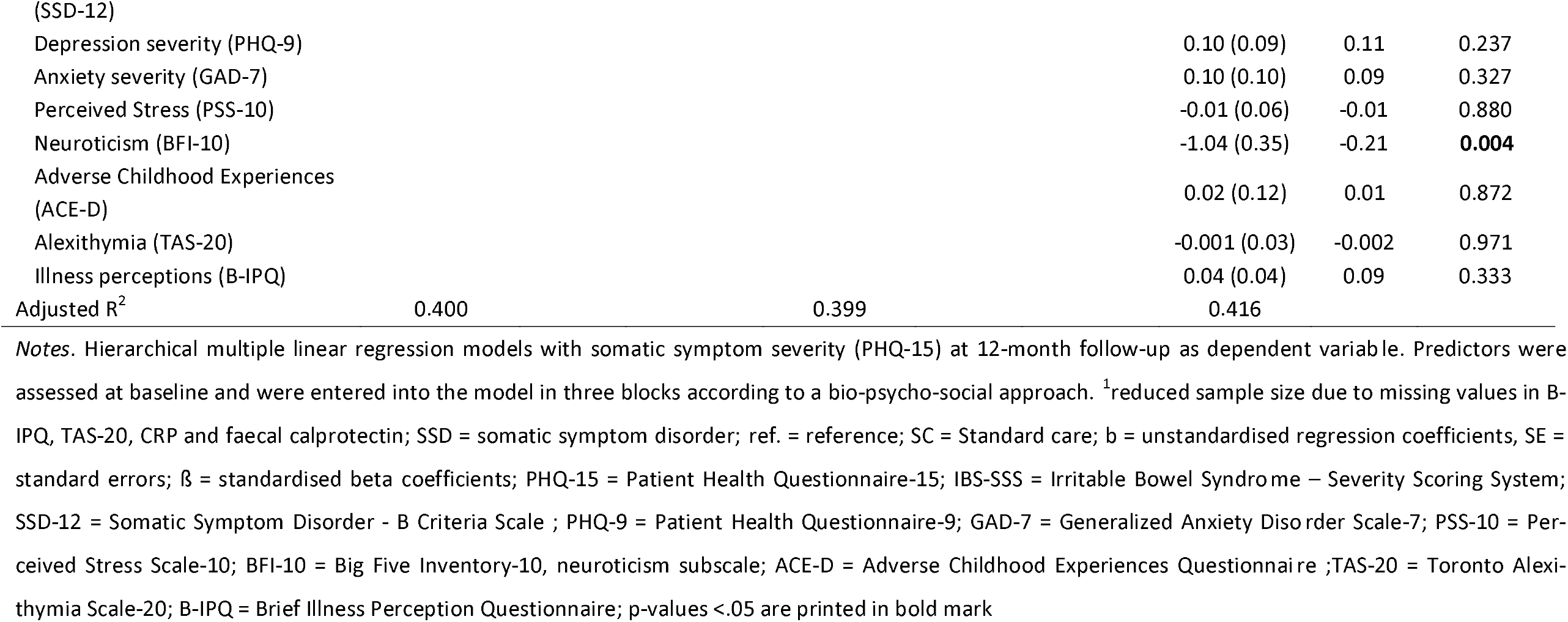
Hierarchical multiple linear regression analysis to test baseline predictors of SSD criterion A assessed via somatic symptom severity (PHQ-15) at 12-month follow-up (N = 196^1^).

**Table 4.**
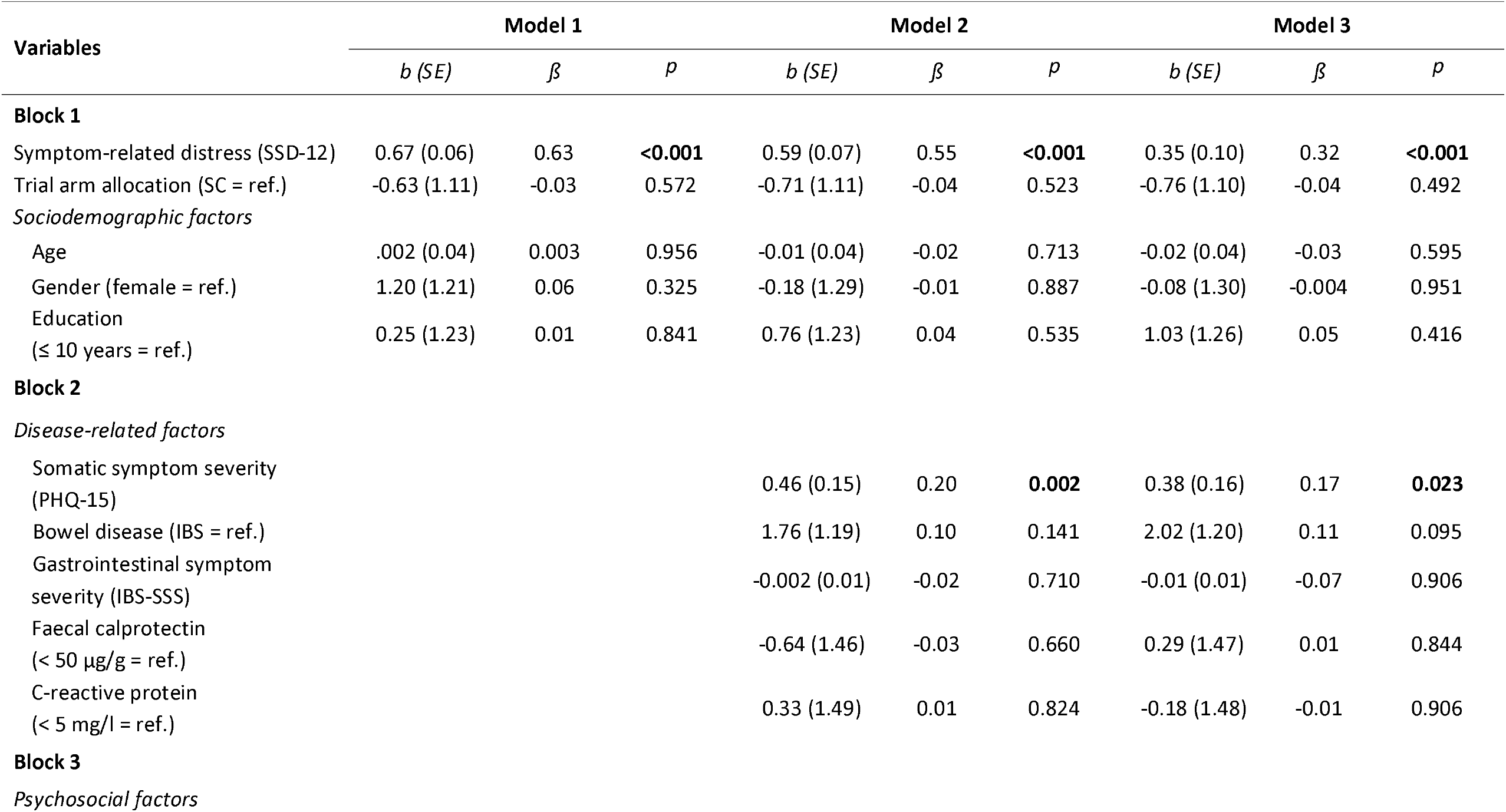
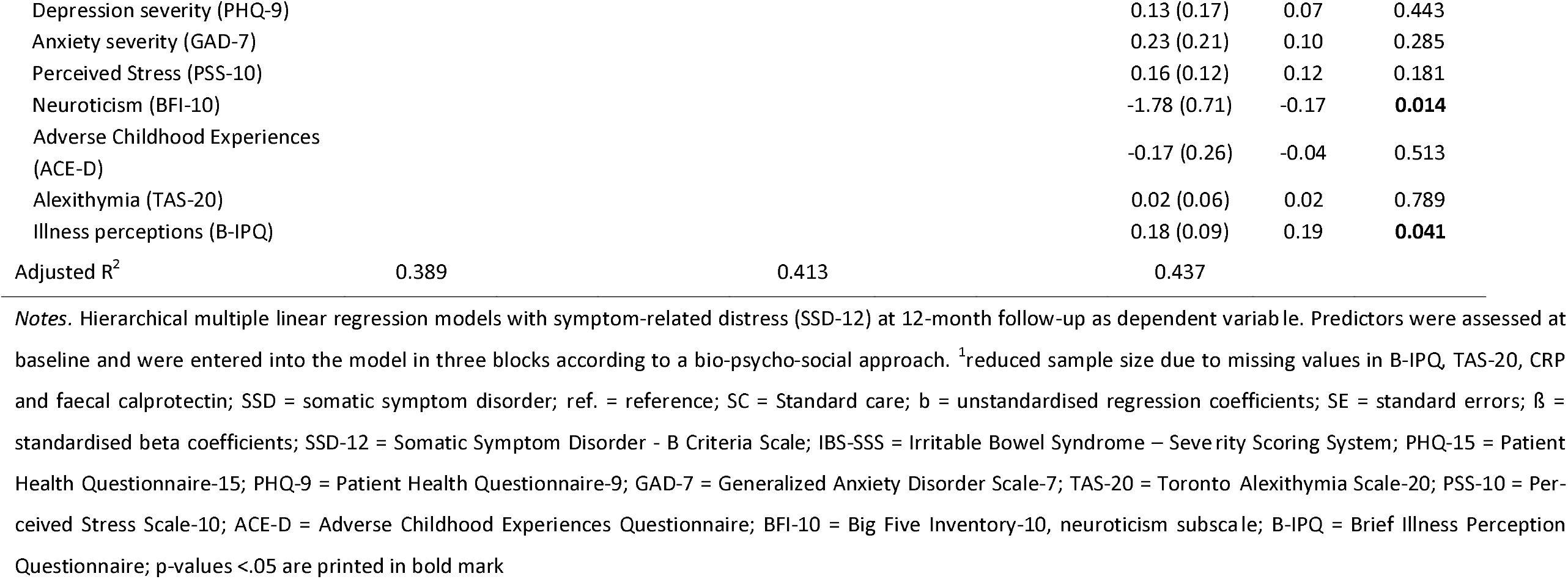
Hierarchical multiple linear regression analysis to test baseline predictors of SSD B criteria assessed via symptom-related distress (SSD-12) at 12-month follow-up (N = 196^1^).

## Discussion

This is the first study to examine the longitudinal course and predictors of SSD in patients with IBS and UC using structured diagnostic interviews. SSD diagnosis was highly prevalent at baseline, affecting over half of patients with IBS and nearly one third of those with UC. Importantly, the results must be interpreted cautiously given the interventional study context, which likely contributed to an overestimated SSD frequency at baseline. Over 12 months, SSD frequency declined substantially in both groups, yet a notable subgroup showed persistent SSD, while around one third remitted and few developed SSD at follow-up. The results extend our previous cross-sectional data and emphasise the relevance of psychosocial factors in predicting SSD in UC and IBS. Notably, no disease-related variable including gastrointestinal symptom severity and inflammatory activity predicted SSD outcome in either IBS or UC. The observed decline contrasts with previous findings from psychosomatic or neurological settings, where SSD rates remained stable or increased (21, 51).

However, these studies investigated the natural course of SSD while the present study used data from an RCT evaluating a psychological intervention. Although no differential treatment effects were detected at the trial’s primary endpoint (23), the setting itself may have influenced outcomes. Participants with greater psychological burden may have been more likely to enroll, increasing the likelihood of regression to the mean (52). Moreover, repeated assessments, regular contact with study staff, and the general effects of trial participation may have encouraged symptom reflection or additional help-seeking, contributing to symptom relief (53). Finally, gastrointestinal symptoms decreased across all trial arms (23), likely reducing symptom-related distress and contributing to lower SSD rates.

The heterogeneous symptom courses in IBS and UC may also account for variability in SSD frequency. Longitudinal studies identified subgroups with stable, improving, or persistently high gastrointestinal symptoms and impaired quality of life in both conditions (54, 55). These studies found that in IBS, gastrointestinal-specific anxiety predicted worsening of symptoms (55), whereas in UC, abdominal pain preceded increases in psychological distress (54). These findings indicate that both cognitive-affective mechanisms and changes in somatic symptom severity can shape symptom-related distress over time, contributing to fluctuations in SSD diagnosis. Consistent with this, low diagnostic stability has been reported for SSD in patients with inflammatory bowel disease: over four years, SSD frequency increased, but none of the baseline cases persisted, while new cases emerged (56). This supports the view that SSD might follow a dynamic rather than stable course, influenced by both symptom intensity and psychological appraisal processes. Patients with IBS seem more vulnerable to SSD, likely due to greater psychological burden, including more negative illness perceptions, higher depression severity, and greater illness-related anxiety compared to those with UC (57-59).

Depression severity and baseline SSD diagnosis predicted SSD at follow-up. This extends our previous cross-sectional findings (19) where illness-related anxiety, neuroticism, and IBS diagnosis were most closely associated with SSD. Longitudinally, depression severity seems to be a central and targetable risk factor for SSD at follow-up aligning with recent meta-analytic findings (18). Clinically, early treatment of depressive symptoms may improve both mood and gastrointestinal symptoms (60), thereby reducing SSD risk. To disentangle risk factors for different aspects of SSD, we also examined predictors of Criteria A and B separately. Criterion A was predicted by baseline somatic symptom severity, female gender, and neuroticism, underscoring the role of stable traits and physical symptom intensity in shaping subsequent symptom severity (61). Criterion B showed a slightly distinct pattern: baseline symptom-related distress, somatic symptom severity, neuroticism and negative illness perceptions emerged as relevant predictors. Inflammatory markers and gastrointestinal symptom severity were not predictive, emphasising the centrality of cognitive and emotional processes in predicting symptom-related distress in both inflammatory and functional bowel disease. Differences between predictors of the categorical SSD diagnosis and the dimensional criteria may reflect distinct underlying mechanisms, but also methodological differences including self-report versus structured interviews with different time frames.

### Clinical implications

The high baseline frequency, subsequent decline, and identified psychosocial predictors of SSD have relevant implications for its diagnosis and management in UC and IBS. Clinicians should systematically consider symptom-related distress, especially in patients with depression or negative illness perceptions, and refer to psychological care if indicated. Notably, even in the absence of marked symptom burden or elevated inflammatory markers, patients may still experience significant psychological distress related to symptoms. In line with the biopsychosocial model of persistent somatic symptoms (62), the findings underscore the clinical value of targeting modifiable risk factors, especially symptom-related distress, maladaptive symptom and illness beliefs, and depressive symptoms in patients with IBS or UC and comorbid SSD (62). Evidence for psychological interventions in SSD is still limited, though first trials indicate that therapist-guided online cognitive behavioral therapy (CBT) addressing e.g., health anxiety, exposure exercises, emotional expression, or CBT strategies can be effective (63-66). CBT has also proven effective in treating somatoform disorders, the former diagnostic category of SSD (67). Future research should test whether interventions tailored to the mechanisms identified in this study improve patient outcomes. Moreover, the results support ntegrated biopsychosocial care models as proposed in current frameworks for persistent somatic symptoms (62). Finally, the findings question the ICD-11 exclusion of BDD in patients with IBS. In this study, SSD was frequent and showed a dynamic course with remitting, persistent, and new cases, indicating that there is overlap between IBS and SSD. Despite overlap, the diagnoses are distinct, defined by different criteria. Excluding BDD in the presence of IBS is insufficiently supported by evidence and may hinder appropriate care. Notably, ICD-11 permits comorbid diagnoses of fibromyalgia and BDD despite some overlapping criteria (68). Revising the current exclusion may enable more accurate clinical decisions and better access to psychological treatment.

### Limitations

Several limitations should be considered when interpreting the findings. First, recruitment within an RCT involving psychological interventions likely selected for individuals with higher psychological burden. The observed decline in SSD frequency may therefore partly reflect treatment-related effects or selection bias, limiting generalisability to unselected populations. Second, patients lost to follow-up reported higher gastrointestinal symptom severity at baseline, suggesting that more severely affected individuals were less likely to complete the study. However, the dropout group was small and otherwise comparable, making systematic bias unlikely. Third, the study combined clinician-rated and self-reported data. While SSD diagnosis was established by structured interview, other variables such as disease activity or symptom severity, relied on self-report and may be prone to recall. The use of validated instruments, inflammatory biomarkers, and structured interviews reduces this risk. Fourth, interpreting SSD B criteria in gastrointestinal conditions remains challenging. Distinguishing intensive coping efforts from excessive symptom-related distress and behaviour is complex and may have influenced diagnosis in both conditions. Future studies should further clarify the operationalisation of SSD B criteria, especially in medical conditions like IBS and UC. Finally, although the longitudinal design is a strength, the 12-month follow-up may not capture the long-term course of SSD in relapsing–remitting conditions like IBS and UC. Longer follow-up in naturalistic settings is needed to better understand SSD trajectories in patients with IBS or UC.

### Conclusion

This study is the first to examine the longitudinal course and biopsychological predictors of diagnostic interview-based SSD in patients with UC or IBS over 12-month within an RCT. SSD was clinically relevant in both groups, with greater vulnerability in IBS compared to UC especially at baseline. SSD rates declined markedly over 12 months, suggesting trial-related effects, but may also indicate a dynamic course. Psychosocial rather than disease-related factors, such as gastrointestinal symptom severity and inflammatory activity, predicted SSD outcomes in both conditions. This underscores the need to identify at-risk patients early and offer tailored interventions addressing modifiable factors such as depressive symptoms and dysfunctional illness perceptions. Future research should examine SSD trajectories in naturalistic settings and test whether targeting the identified mechanisms improve clinical outcomes in patients with IBS or UC and comorbid SSD.

## Data Availability

Data can be requested from the corresponding author upon reasonable request. Data use and requests underlie data protection and the publication policy of the Research Unit SOMACROSS

## Acknowledgment

The authors wish to thank all patients for their participation in the SOMA.GUT-RCT. Furthermore, the authors wish to thank the psychological research assistants Sophie Schmitz, Xenia Schell, Serhat Karayilan and the medical research assistants Kirsten Hammer, Sarika Schumacher and Oumayma Chbani for conducting the SSD interviews with all patients enrolled in the study.

## Statement of Ethics

The study was approved by the Ethics Committee of the Hamburg Medical Association (reference number: 2020-10198-BO-ff).

## Declaration of Generative AI and AI-assisted technologies in the writing process

During the preparation of this work, the authors used ChatGPT in order to improve the wording. After using this tool, the authors reviewed and edited the content as needed and take full responsibility for the content of the publication.

## Author contributions

Conceptiualization: LP KM BL. Methodology: LP BL KM. Software: LP KM. Validation: LP BL KM. Formal analysis: LP BL KM. Investigation: LP AM SH. Data curation: LP KM AM SH. Writing – Original Draft: LP. Writing – Review & Editing: KM BL AWL AM SH. Visualization: LP. Supervision: KM BL. Project administration: KM BL. Funding acquisition: BL AWL

