## Supplement 1 for "Course and predictors of somatic symptom disorder in irritable bowel syndrome and ulcerative colitis: A longitudinal analysis from the SOMA.GUT-RCT"

*Sociodemographic and disease-related characteristics of the study sample and the dropout group*

| **Variables** | **Study sample**  *n* = 213  *M* (*SD*)/*n* (%) | **Dropout group**  *n* = 23  *M* (*SD*)/*n* (%) | ***t/χ2*** | ***df*** | ***p*** |
| --- | --- | --- | --- | --- | --- |
| **SSD diagnosis at baseline** | 90 (42.3) | 8 (34.8) | 0.48 | 1 | 0.490 |
| **Group allocation within the SOMA.GUT-RCT** |  |  |  |  |  |
| Standard Care (SC) | 69 (32.4) | 12 (52.2) | 4.43 | 2 | 0.109 |
| GUT.EXPECT+SC | 68 (31.9) | 7 (30.4) |  |  |  |
| GUT.SUPPORT+SC | 76 (35.7) | 4 (17.4) |  |  |  |
| **Age** |  |  |  |  |  |
| Years | 40.51 (13.98) | 36.72 (11.33) | -1.26 | 234 | 0.210 |
| **Gender** |  |  |  |  |  |
| Male | 54 (25.4) | 6 (26.1) | 0.22 | 1 | 0.896 |
| Female | 157 (73.7) | 17 (73.9) |  |  |  |
| Diverse | 2 (0.9) | / |  |  |  |
| **Education** |  |  |  |  |  |
| ≤ 10 years | 52 (24.4) | 4 (17.4) | 1.82 | 2 | 0.403 |
| High school diploma | 64 (30.0) | 10 (43.5) |  |  |  |
| University graduation | 97 (45.5) | 9 (39.1) |  |  |  |
| **Employment** |  |  |  |  |  |
| Full-time | 91 (42.7) | 10 (43.5) | 1.14 | 4 | 0.889 |
| Part-time | 54 (25.4) | 5 (21.7) |  |  |  |
| Not employed | 6 (2.8) | 1 (4.3) |  |  |  |
| Retired | 19 (8.9) | 1 (4.3) |  |  |  |
| Other (e.g. parental leave, in education) | 43 (20.2) | 6 (26.1) |  |  |  |
| **Marital status** |  |  |  |  |  |
| Single | 108 (50.7) | 12 (52.2) | 2.02 | 3 | 0.568 |
| Married | 84 (39.4) | 8 (34.8) |  |  |  |
| Divorced/separate living | 19 (8.9) | 2 (8.7) |  |  |  |
| Widowed | 2 (0.9) | 1 (4.3) |  |  |  |
| **Bowel disease** |  |  |  |  |  |
| UC | 110 (51.6) | 15 (65.2) | 1.54 | 1 | 0.215 |
| IBS | 103 (48.4) | 8 (34.8) |  |  |  |
| **Disease duration in years** | 8.96 (9.19) | 8.82 (7.22) | -0.07 | 234 | 944 |
| **Symptom duration in years** | 13.39 (11.56) | 12.45 (10.76) | -0.36 | 233 | 0.718 |
| **Gastrointestinal symptom severity (IBS-SSS)** | 264.32 (92.89) | 311.30 (92.26) | 2.31 | 234 | **0.022** |
| **Flare-up yes (UC only)** | 45 (40.9) | 7 (46.7) | 0.18 | 1 | 0.671 |
| **Disease activity**  **(SSCAI, UC only)** | 5.07 (2.69) | 5.20 (2.60) | 0.17 | 123 | 0.863 |
| **Faecal calprotectin^1^** |  |  |  |  |  |
| µg/g | 147.57 (597.12) | 231.89 (557.69) | 0.58 | 224 | 0.564 |
| % with ≥ 50 µg/g | 41 (19.2) | 6 (26.1) | 0.61 | 1 | 0.435 |
| **C-reactive protein (CRP)^2^** |  |  |  |  |  |
| mg/l | 3.47 (4.16) | 3.0 (3.11) | -0.46 | 222 | 0.643 |
| % with ≥ 5 mg/l | 32 (15.0) | 2 (8.7) | 0.67 | 1 | 0.412 |

*Notes.* UC = ulcerative colitis; IBS = irritable bowel syndrome; SSD = somatic symptom disorder; SC = Standard Care; GUT.EXPECT = expectation management intervention in the SOMA.GUT-RCT; GUT.SUPPORT = unspecific supportive intervention in the SOMA.GUT-RCT = ; IBS-SSS = Irritable Bowel Syndrome – Severity Scoring System; SCCAI = Simple Clinical Colitis Activity Index*;* ^1^ n = 5 missing values, values under the detection limit were imputed with half-detection limit 2.5 µg/g; ^2^ n = 7 missing values, values under the detection limit were imputed with half-detection limit 2.0 mg/l, *p*-values <.05 are printed in bold mark.
