## Supplement 2 for "Course and predictors of somatic symptom disorder in irritable bowel syndrome and ulcerative colitis: A longitudinal analysis from the SOMA.GUT-RCT"

### Slide 1
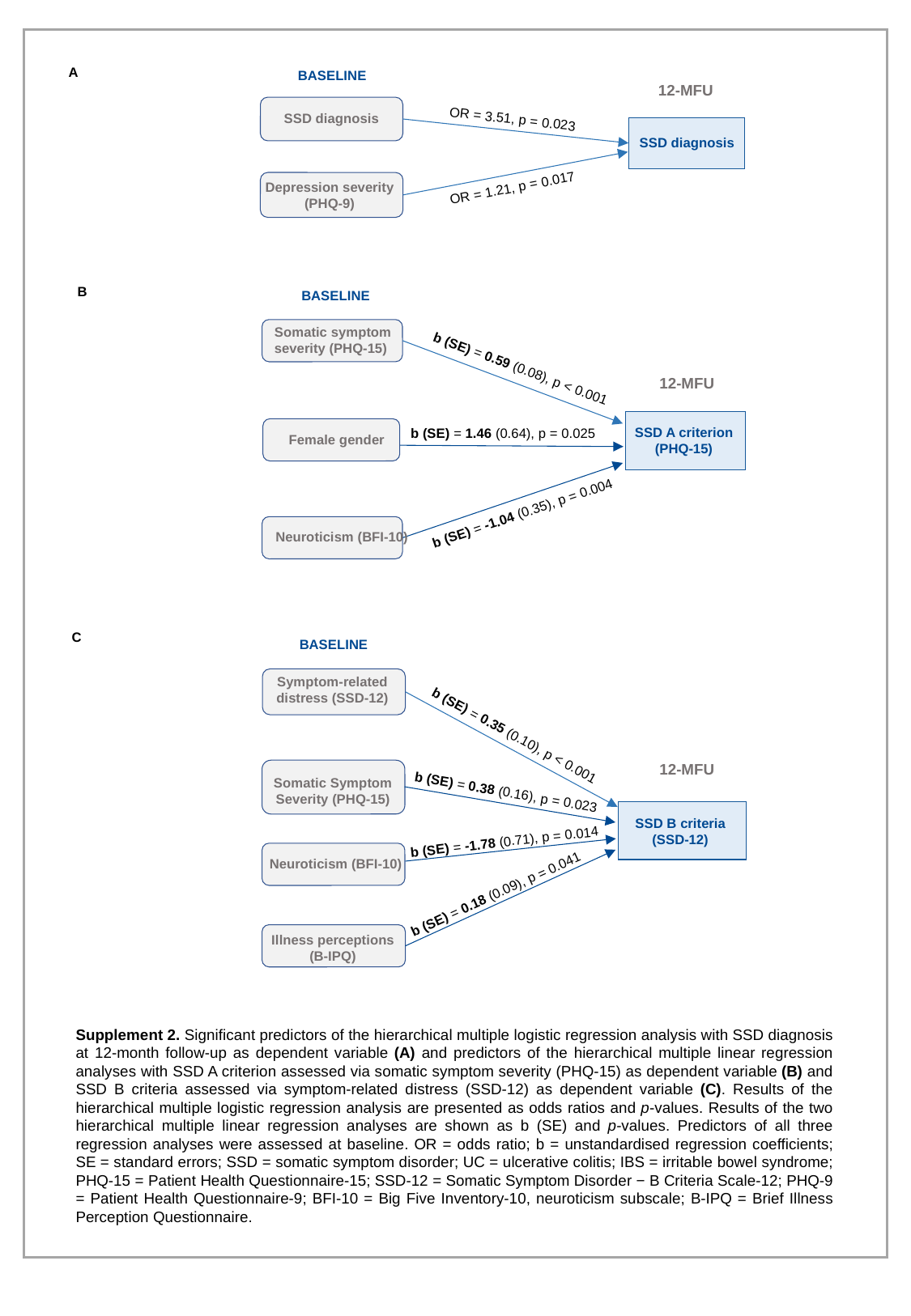

A
BASELINE
SSD diagnosis
OR = 3.51, p = 0.023
SSD diagnosis
Depression severity (PHQ-9)
OR = 1.21, p = 0.017
12-MFU
B
BASELINE
Somatic symptom severity (PHQ-15)
SSD A criterion (PHQ-15)
b (SE) = 1.46 (0.64), p = 0.025
Female gender
b (SE) = -1.04 (0.35), p = 0.004
Neuroticism (BFI-10)
b (SE) = 0.59 (0.08), p < 0.001
12-MFU
C
BASELINE
Symptom-related distress (SSD-12)
b (SE) = 0.35 (0.10), p < 0.001
SSD B criteria (SSD-12)
b (SE) = -1.78 (0.71), p = 0.014
Illness perceptions (B-IPQ)
Neuroticism (BFI-10)
b (SE) = 0.18 (0.09), p = 0.041
12-MFU
Somatic Symptom Severity (PHQ-15)
b (SE) = 0.38 (0.16), p = 0.023
Supplement 2. Significant predictors of the hierarchical multiple logistic regression analysis with SSD diagnosis at 12-month follow-up as dependent variable (A) and predictors of the hierarchical multiple linear regression analyses with SSD A criterion assessed via somatic symptom severity (PHQ-15) as dependent variable (B) and SSD B criteria assessed via symptom-related distress (SSD-12) as dependent variable (C). Results of the hierarchical multiple logistic regression analysis are presented as odds ratios and p-values. Results of the two hierarchical multiple linear regression analyses are shown as b (SE) and p-values. Predictors of all three regression analyses were assessed at baseline. OR = odds ratio; b = unstandardised regression coefficients; SE = standard errors; SSD = somatic symptom disorder; UC = ulcerative colitis; IBS = irritable bowel syndrome; PHQ-15 = Patient Health Questionnaire-15; SSD-12 = Somatic Symptom Disorder − B Criteria Scale-12; PHQ-9 = Patient Health Questionnaire-9; BFI-10 = Big Five Inventory-10, neuroticism subscale; B-IPQ = Brief Illness Perception Questionnaire.
